# Clinical Outcomes of Patients with COVID-19 and Chronic Inflammatory and Autoimmune Rheumatic Diseases: A Multicentric Matched-Cohort Study

**DOI:** 10.1101/2020.06.18.20133645

**Authors:** José L. Pablos, María Galindo, Loreto Carmona, Miriam Retuerto, Ana Lledó, Ricardo Blanco, Miguel A. González-Gay, David Martinez-Lopez, Isabel Castrejón, José M. Álvaro-Gracia, David Fernández-Fernández, Antonio Mera-Varela, Sara Manrique-Arija, Natalia Mena-Vázquez, Antonio Fernández-Nebro, RIER investigators group

**Author notes:** Correspondence to José L. Pablos, Servicio de Reumatología, Hospital 12 de Octubre; 28041 Madrid, Spain. RIER investigators group: Rodrigo Aguirre, Álvaro Seijas-López, Francisco J Blanco, (Servicio de Reumatología, INIBIC-Complejo Hospitalario Universitario A Coruña, Universidad de A Coruña, A Coruña, Spain); Patricia Carreira, María Martín-López (Servicio de Reumatología, Instituto de Investigación Hospital 12 de Octubre (imas12), Universidad Complutense de Madrid, Madrid, Spain); Antonio Gonzalez (Instituto de Investigación Sanitaria de Santiago (IDIS), Santiago de Compostela, Spain); Amaya Puig-Kröger, Luis Salas (Instituto de Investigación Sanitaria Gregorio Marañón (IiSGM), Madrid, Spain).

## Abstract

**Background:** The impact of inflammatory rheumatic diseases on COVID-19 severity is poorly known. Here we compare the outcomes of a cohort of rheumatic patients with a matched control cohort to identify potential risk factors for severe illness.

**Methods:** In this comparative cohort study, we identified hospital PCR+ COVID-19 rheumatic patients with chronic inflammatory arthritis (IA) or autoimmune/immunomediated diseases (AI/IMID). Non-rheumatic controls were randomly sampled 1:1, and matched by age, sex, and PCR date. The main outcome was severe COVID-19, defined as death, invasive ventilation, ICU admission, or serious complications. We assessed the association between the outcome and potential prognostic variables, adjusted by COVID treatment, using logistic regression.

**Results:** The cohorts were composed of 456 rheumatic and non-rheumatic patients, in equal numbers. Mean age was 63 [IQR 53-78] and male sex 41% in both cohorts. Rheumatic diseases were IA (60%) and AI/IMID (40%). Most patients (74%) had been hospitalised, and the risk of severe COVID was 31.6% in the rheumatic and 28.1% in the non-rheumatic cohort. Ageing, male sex and previous comorbidity (obesity, diabetes, hypertension, cardiovascular, or lung disease) increased the risk in the rheumatic cohort by bivariate analysis. In logistic regression analysis, independent factors associated with severe COVID were increased age (OR 5.31; CI 3.14-8.95), male sex (2.13; CI 1.35-3.36) and having an AI/IMID (OR 1.98; CI 1.15-3.41).

**Conclusion:** In patients with chronic inflammatory rheumatic diseases aging, sex and having an AI/IMID but not IA nor previous immunosuppressive therapies were associated with severe COVID-19.

## INTRODUCTION

The clinical spectrum of SARS-CoV-2 infection is quite broad, ranging from asymptomatic to life-threatening or fatal disease. Different factors have been associated to poor prognosis, including older age, gender, and pre-existing comorbidities such as diabetes, hypertension and lung and cardiovascular disease The risk for severe COVID-19 has been associated with older age, gender, and pre-existing morbidities such as diabetes, lung and cardiovascular disease including hypertension [1-3]. Immune-mediated conditions and immunosuppressive therapies increase the susceptibility to viral and bacterial infections and therefore, understanding how COVID-19 impacts on these patients is an urgent need [4-6].

Since the severity of COVID-19 is associated with a hyperinflammatory process, it is also of particular interest to investigate how pre-existing immune disturbances or the previous use of immunosuppressive agents influence COVID-19 expression [7]. We have previously reported an increased prevalence of hospital attended COVID-19 in patients with AI/IMID, and specifically in those on ts/dDMARD therapy, compared to a reference population, reflecting either increased risk of infection or increased severity [8]. In the larger COVID-19 series, neither AI/IMID nor immunosuppressive therapies are represented, but incompleteness of information on these specific factors is possible [1-3]. In recent report of a small cohort of hospitalized rheumatic patients, a higher need for mechanical ventilation compared to non-rheumatic controls has been reported [9]. A global registry of rheumatic patients found corticosteroids (CS) but not other therapies to a higher risk for hospitalization [10]. In patients with inflammatory bowel disease, CS but not anti-TNF-α drugs independently increase the risk of severe disease [11]. Other immunosuppressed patients, as solid organ transplanted, have more severe COVID-19, however the role of age or comorbidities, and the lack of controls do not permit to draw definitive conclusions [12,13].

An additional concern among rheumatologists is that in most chronic inflammatory rheumatic diseases, clinical and subclinical metabolic and cardiovascular comorbidity is increased. which may also put these patients at higher risk of poor outcomes [14, 15]. It is therefore necessary for contingency prevention plans, to identify vulnerable patients and specific features at high risk requiring special vigilance or management.

We undertook a multicentric comparative cohort study to investigate the relationship between underlying rheumatic disease and COVID-19 outcomes and to identify specific risk factors associated to poor outcomes.

## PATIENTS AND METHODS

We performed a retrospective observational matched-cohort study from the databases of five reference centres pertaining to a public research network for the investigation of inflammation and rheumatic diseases (RIER, https://red-rier.org/). Each of the included centres have accessibility to updated medical records ID lists of adult patients under follow-up in rheumatology departments, and were reference centres for microbiology, where all SARS-CoV-2 PCR diagnostic tests in the covered population were performed. Patients’ medical record IDs were matched against central SARS-CoV-2+ PCR hospital registers up to April 17^th^, just after the incidence peak of SARS-CoV-2 infection had been reached in Spain [https://cnecovid.isciii.es/covid19]. Electronic medical records were reviewed to confirm COVID-19 diagnosis and to obtain clinical data. Since at that time, availability of CoV-2 PCR testing was limited due to shortages, these registries only include patients attending referral hospitals, and exclude the less severe community cases that did not require hospitalization nor referral to hospitals emergency departments.

The rheumatology cohort included all adult patients diagnosed of chronic inflammatory arthritis (IA), including rheumatoid arthritis, psoriatic arthritis (PsA) and spondyloarthritis (SpA); AI/IMID, including systemic lupus erythematosus (SLE), Sjögren’s syndrome (SS), systemic sclerosis (SSc), polymyalgia rheumatica (PMR), vasculitides etc (Supplementary Table S1) with a PCR+ COVID-19 diagnosis. The control cohort was assembled from the Microbiology databases of the participating centres matched on a 1:1 basis with the rheumatic cohort on the date of COVID-19 diagnosis (“index date”), sex, and age, and blinded to outcome or other variables. In this control cohort, patients with IA/IMID were excluded.

### Variables and measurements

We collected the following data from the electronic health record to descrive COVID-19 evolution: evidence of pneumonia by plain X-ray, respiratory insufficiency, oxygen necessities (collected as ordinal variable ranging from 0 “no external oxygen required”, to 1 “oxygen by nasal cannula”, 2 “reservoir”, 3 “non-invasive ventilation”, and 4 “tracheal intubation”), serious life threatening complications (myocarditis or heart failure, encephalopathy, thrombosis, kidney failure or septic shock), duration of admission, and exitus. Laboratory data were also collected at baseline and at peak levels for the following variables: C-reactive protein (CRP), interleukin-6 (IL-6), lymphocyte counts, D-Dimer, lactate dehydrogenase (LDH), and ferritin.

The primary outcome was a composite outcome, ‘severe COVID’, including death, ICU admission, intratracheal intubation, or serious COVID-19 complications. Factors studied in relation to the outcome were those common to all COVID patients, such as age (with a cut-off at 60 years), male sex, cardiovascular disease, obesity, diabetes, hypertension, and lung disease. Specific factors for rheumatic diseases included diagnostic group, disease duration, treatments—such as glucocorticoids (GC), conventional synthetic disease-modifying antirheumatic drugs csDMARD, other immunosuppressants (including azathioprine, cyclophosphamide, mofetil mycophenolate, and calcineurin inhibitors), or ts/bDMARD, including jakinibs (tofacitinib or baricitinib), or any biological agent (TNF-α, IL-1, IL-6 or IL23/17 antagonists, abatacept or rituximab). COVID-19 treatment was also collected and treated as potentially confounding covariate. The most commonly used treatments were hydroxychloroquine (HCQ), antivirals (lopinavir/ritonavir and remdesivir), GC and anti-cytokines. We used summary statistics to describe the cohorts, and t tests, chi-square, Fisher’s exact, and log rank tests to refute hypothetical differences between them. For time to variables we used May 15^th^ as censor date.

We then estimated the risk to develop severe COVID in each cohort, in terms of point estimates and 95% confidence intervals (CI), risk difference, risk ratio, and attributable fractions for the rheumatic and total population. The relative risk of prognostic factors was estimated, and the hypothesis of an effect modification of having an AI/IMID rheumatic disease tested with the Mantel-Haenszel method.

Subsequently we run bivariable and multivariable logistic regression models to assess the association rheumatic diseases and severe COVID in detail, where the composite outcome was the dependent variable. We used several approaches to building the models: (1) using the *allsets* command, (2) automatic backward stepwise starting with a full model with all variables with a p-value < 0.25 in the bivariable, and (3) a manual stepwise method, keeping cohort and confounding variables in the model. The best model was selected on the basis of the AIC and BIC criteria, and the area under the ROC curve and predictive capacity of the best model estimated. All analyses were done in Stata 12 (College Station, Texas, USA).

All data were anonymised, and the study was approved by the Ethics Committee of Hospital 12 de Octubre Hospital in April 3^th^, with ref N° CEIm: 20/160.

## RESULTS

The total sample was 456, evenly distributed into 228 patients per cohort. The diagnoses of rheumatic patients were IA (n=136, 60%): RA (n=65, 29%), SpA (n=35, 16%), PsA (n=36, 15%), and AI/IMID (92, 40%) as detailed in Table S1. The mean duration of the rheumatic disease was 10 years (SD 8.3) with no differences across diseases.

Table 1 shows a description of both cohorts. These were matched in terms of age and sex, and well balanced regarding most other variables. However, clinician reported obesity, and cardiovascular disease were more frequent among rheumatic patients versus controls.

**Table 1.**
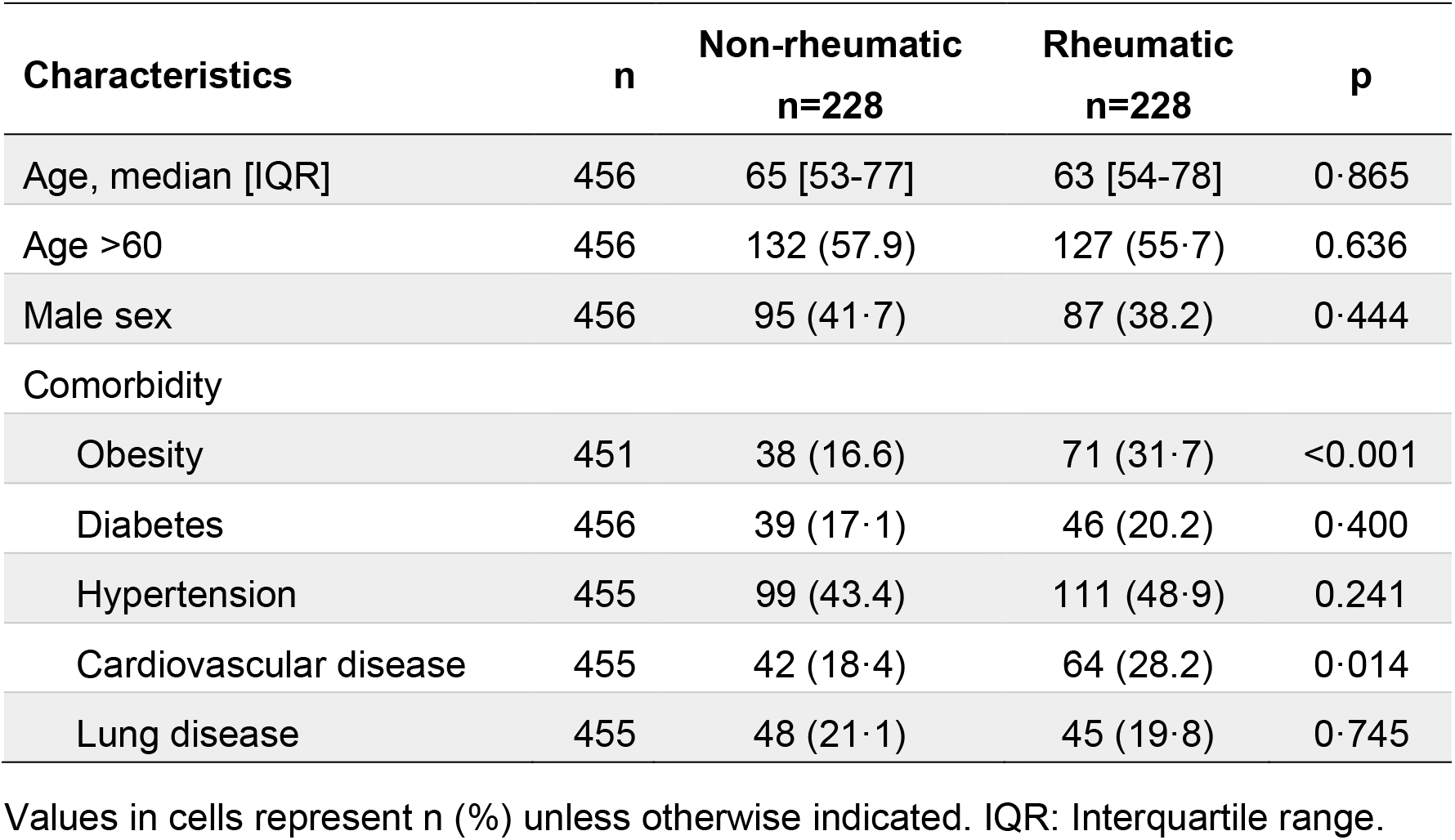
Description of the cohorts compared.

Regarding treatments used by the rheumatic patients that could predispose them to infection, most patients were on csDMARDs (57%), followed by glucocorticoids (40%), biologic agents (23%), mostly TNF-α antagonists, and 12% on other immunosuppressants (Table 2). No patient in the non-rheumatic cohort was taking any of these drugs, except for a patient who was taking glucocorticoids at a dose of 5 mg/d for other reasons.

**Table 2.** Drugs used by the rheumatic cohort at the time of cohort entry.

| **Treatment** | **n (%)** |
| --- | --- |
| **Glucocorticoids** | 91 (40.1) |
| Dose*, m ± SD when taken | 9.9 ± 11.5 |
| **csDMARD** | 129 (56.6) |
| Methotrexate | 64 (28.1) |
| Antimalarial drugs | 28 (12.4) |
| Leflunomide | 20 (8.9) |
| Sulphasalazine | 17 (7.5) |
| **Other immunosuppressants** | 28 (12.3) |
| Mofetil mycophenolate | 12 (5.3) |
| Azathioprine | 7 (3.1) |
| Cyclophosphamide | 2 (0.8) |
| Calcineurin inhibitors | 7 (3.1) |
| **ts/bDMARD** | 53 (23.2) |
| TNF-α antagonists | 35 (15.4) |
| Rituximab | 5 (2.2) |
| IL-17/IL23 antagonists | 4 (1.8) |
| Abatacept | 3 (1.3) |
| Tocilizumab | 2 (0.8) |
| Sarilumab | 1 (0.4) |
| Tofacitinib | 3 (1.3) |

\*In mg/d of prednisone equivalents.

The evolution of the COVID-19 disease and its comparison between cohorts is described in Table 3. The bivariable analysis shows a larger proportion of radiographic pneumonia in the non-rheumatic cohort and a non-statistically larger proportion of myocarditis and higher peak serum creatinine in the rheumatic cohort. All other variables measured were similar between groups, which were also treated with similar drugs, with the exception of a non-statistically significant larger use of azithromycin in the non-rheumatic cohort.

**Table 3.**
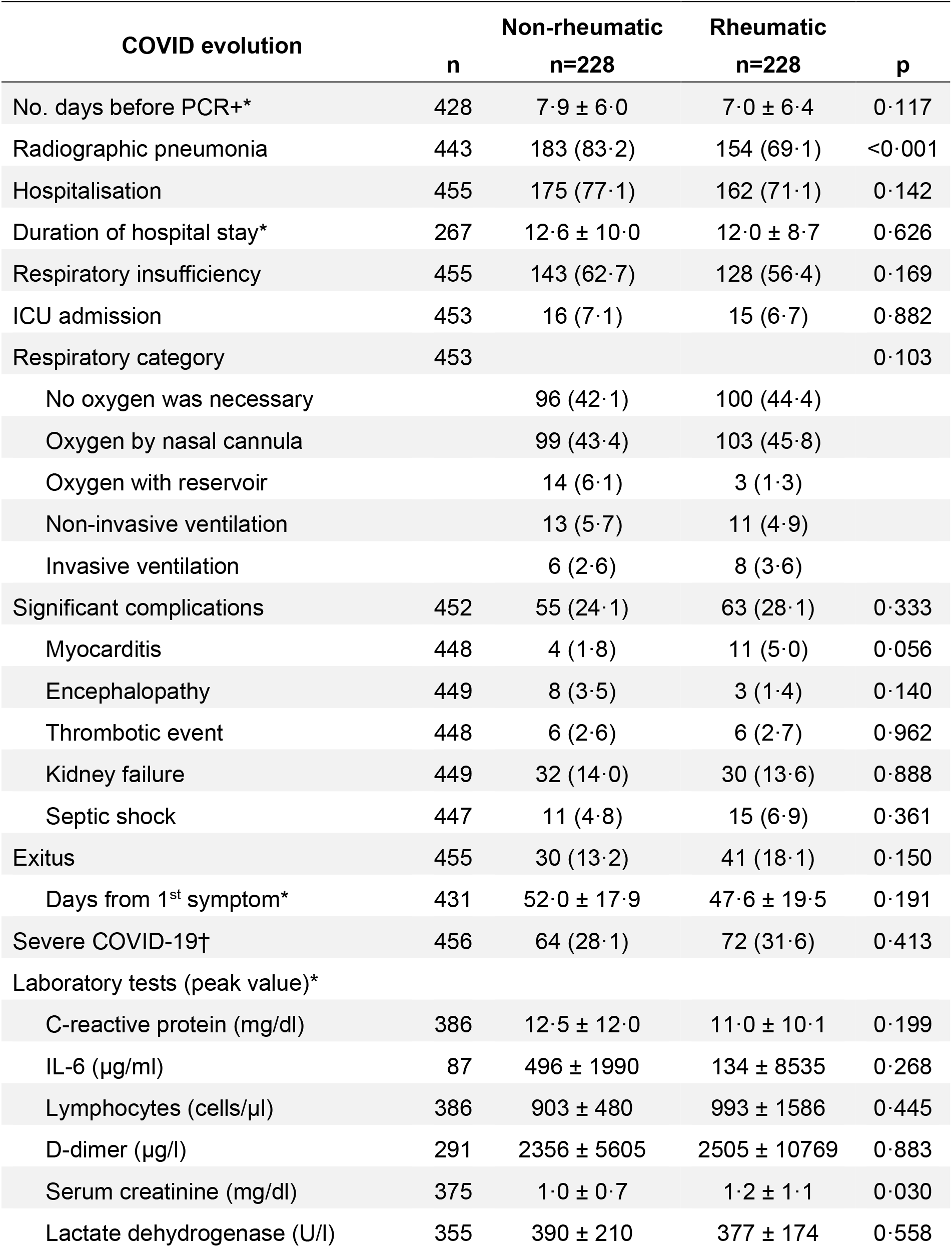
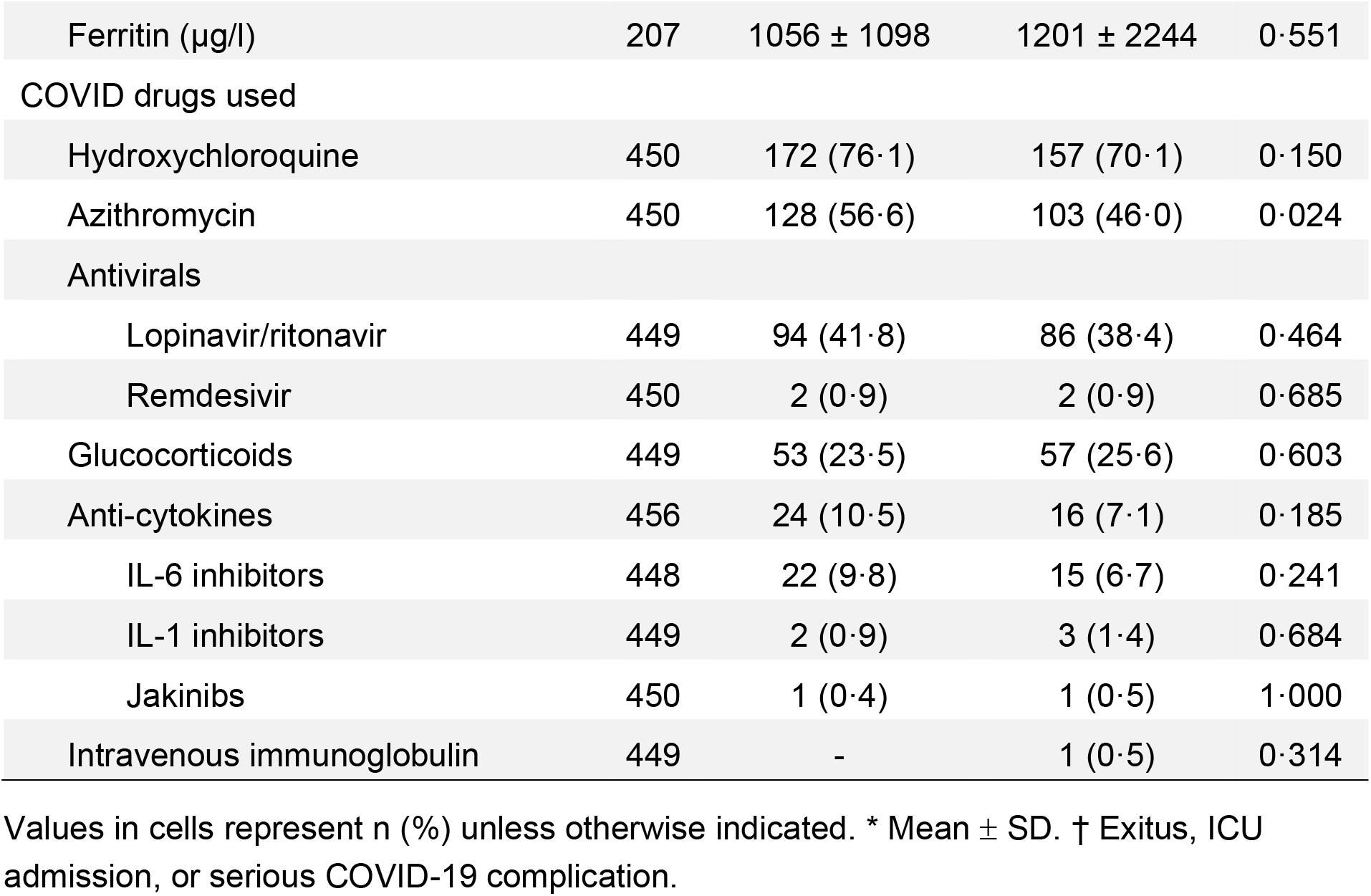
Description of evolution and therapy of COVID-19 in the compared cohorts.

The risk of a severe COVID was 28.1% in the non-rheumatic cohort and of 31.6% in the rheumatic cohort, i.e., a risk difference of 3.5% (95% CI: −4.9% to 11.9%), a risk ratio of 1.13 (95% CI 0.84 to 1.49), an attributable fraction of the exposed of 11.1%, and in the population of 5.9% (*p* = 0.413).

Table 4 shows the relative risk of variables common to both cohorts, by cohort. Age ≥60 and all comorbid variables were associated with outcome in the rheumatic cohort, but only age, hypertension and lung disease not in the non-rheumatic cohort. No clear effect modification of the cohort on the associations was present, as by the results of the homogeneity test.

**Table 4.** Analysis of individual risk factors for poor outcome; total and by cohort.

| Variable | Relative risk (95% confidence interval) |  | *p* * |
| --- | --- | --- | --- |
|  | Non-rheumatic cohort | Rheumatic cohort |  |
| Age over 60 | **3.70 (1.99 - 6.93)** | **4.04 (2.30 - 7.08)** | 0.841 |
| Male sex | **2.16 (1.39 - 3.35)** | **1.58 (1.09 - 2.29)** | 0.286 |
| Obesity | 1.22 (0.72 - 2.06) | **1.62 (1.10 - 2.36)** | 0.393 |
| Diabetes | 0.95 (0.53 - 1.70) | **1.93 (1.34 - 2.79)** | 0.038 |
| Hypertension | **1.64 (1.07 - 2.53)** | **2.27 (1.49 - 3.46)** | 0.290 |
| CV disease | 1.44 (0.90 - 2.33) | **2.92 (2.04 - 4.17)** | 0.020 |
| Lung disease | **1.57 (1.00 - 2.46)** | **1.74 (1.19 - 2.55)** | 0.723 |

\* From a Mantel-Haenszel test of homogeneity. If $p < 0.01$ the cohort of origin is modifying the effect. In bold statistically significant associations with outcome. Abbreviation: CV, cardiovascular.

The results of the bi- and multivariable logistic regression analysis are shown in Table 5. Variables with very low observations were not analysed or combined into meaningful categories. The best model was the stepwise automatic one, and its results are shown in the right part of the table and in Figure 1. The model correctly classified 72.23% of the patients (AUC = 0.753).

**Table 5.** Association of risk factors with poor outcome in COVID-19

| Factors | OR (95% CI)<br>Bivariable | p | OR (95% CI)<br>Multivariable | p |
| --- | --- | --- | --- | --- |
| Rheumatic disease | 1.29 (0.86 - 1.93) | 0.218 |  |  |
| AI/IMID | 1.64 (1.02 - 2.66) | 0.042 | 1.98 (1.15 - 3.41) | 0.014 |
| Chronic IA | 0.90 (0.58 - 1.41) | 0.659 |  |  |
| Age > 60 | 6.06 (3.65 - 10.06) | <0.001 | 5.31 (3.14 - 8.95) | <0.001 |
| Male sex | 2.34 (1.55 - 3.53) | <0.001 | 2.13 (1.35 - 3.36) | 0.001 |
| Comorbidity |  |  |  |  |
| Obesity | 1.78 (1.13 - 2.81) | 0.013 |  |  |
| Diabetes | 1.81 (1.11 - 2.95) | 0.018 |  |  |
| Hypertension | 2.60 (1.72 - 3.94) | <0.001 |  |  |
| CV disease | 3.49 (2.21 - 5.51) | <0.001 |  |  |
| Lung disease | 2.15 (1.34 - 3.45) | 0.001 |  |  |
| Medication |  |  |  |  |
| Glucocorticoids<br>(any dose) | 2.20 (1.36 - 3.54) | 0.001 |  |  |
| HCQ | 1.15 (0.51 - 2.62) | 0.733 |  |  |
| csDMARDs | 1.04 (0.64 - 1.72) | 0.864 |  |  |
| b/tsDMARDs | 0.45 (0.21 - 0.96) | 0.039 |  |  |
| Other IS | 0.99 (0.40 - 2.44) | 0.981 |  |  |
| COVID drugs used |  |  |  |  |
| HCQ | 2.26 (1.35 - 3.79) | 0.002 |  |  |
| Antivirals | 2.37 (1.58 - 3.59) | <0.001 | 2.05 (1.30 - 3.23) | <0.001 |

Abbreviations: OR, odds ratio; CI, confidence interval; CTD, connective tissue disease; CV, cardiovascular; csDMARD, conventional synthetic disease-modifying drugs; ts/bDMARD, targeted synthetic or biological disease-modifying drugs; IS, immunosuppressants; HCQ, hydroxychloroquine

**Figure 1.**
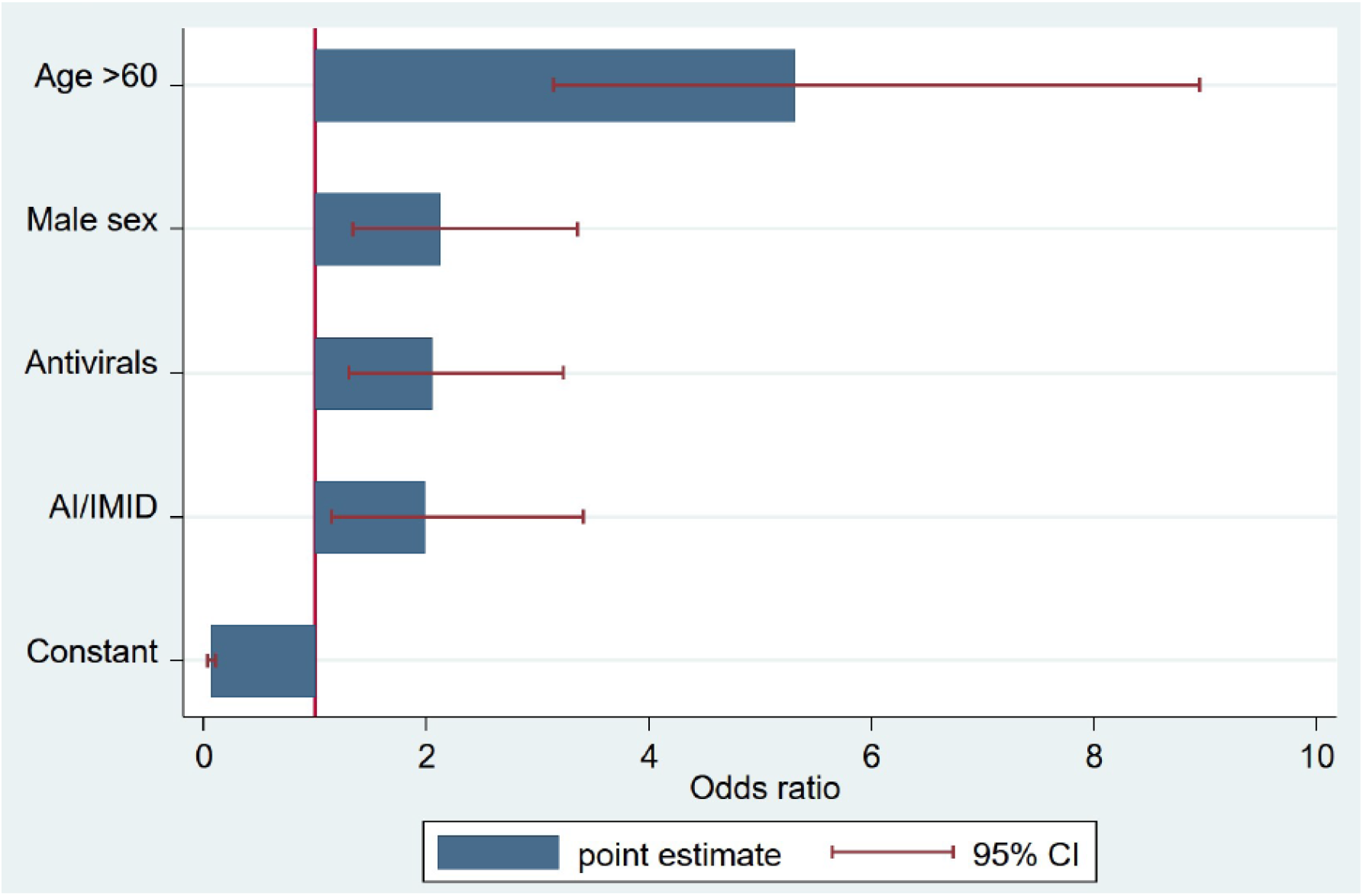
Odds ratios with 95% confidence intervals of the best fitted model to predict ‘severe COVID’.

An independent association between AI/IMID (OR 1.98; CI 1.15-3.41), age (OR 5.31; CI 3.14 - 8.95), and male sex (OR 2.13; OR 1.35 - 3.36) with higher risks for the composite severe COVID-19 outcome was found, whereas all other factors IA, comorbidities and active anti-rheumatic therapies were not confirmed in the multivariable adjusted analysis. A higher use of antivirals also remained associated to severity.

## DISCUSSION

In this matched-cohort study, we show that among hospital patients with chronic inflammatory rheumatic diseases, having a systemic AI/IMID but not an IA is an independent risk factor for poor COVID outcomes including death, invasive ventilation, ICU admission or serious complications. Comorbidities known to associate with severe COVID-19 in the general population, are also associated with greater risk to these patients by bivariable analyses [1-3, 14]. This observation is of particular interest, because some of them as cardiovascular disease, obesity and metabolic syndrome, are also associated with inflammatory disease as also shown in our cohort [15,16]. However, there was no statistically significant association between these morbidities and severity by multivariable analysis, suggesting collinearity with ageing and inflammatory disease.

Our data are in agreement with a previous study in a smaller COVID-19 hospitalized cohort, which also identified a higher odds of intensive care admission/mechanical ventilation among hospitalised patients with rheumatic disease versus matched controls [9]. In contrast with ours, this study combined IA and AI/IMID patients. Our data illustrate how these groups may carry a different risk for severe COVID-19. In the present study, only AI/IMID patients show a higher risk whereas IA patients do not. Whether specific diagnostics among these groups may have a different risk cannot be ruled-out due to statistical power limitation. In our previous prevalence analysis, hospital cases were more prevalent among SpA but not RA or PSA groups, and in AI/IMID groups but not SLE patients compared to the reference population [8]. Further analyses would be needed to evaluate the severity of specific rheumatic diseases.

Concerning previous use of therapies by rheumatic patients, the use of glucocorticoids was associated with poorer outcome by bivariable analysis, whereas no substantial risk was detected neither for traditional immunosuppressants nor csDMARDS (methothrexate and leflunomide), nor for bDMARD (mostly anti-TNF-α). Interestingly, the use of ts/bDMARD was associated in the bivariable with lower odds of complications. They did not make it into the final models probably because of the collinearity with other variables and not being the full rheumatic sample included, thus encountering problems of statistical power.

The potential therapeutic effect of anti-cytokine biologicals and Jakinibs on COVID-19 is being tested through numerous observational and randomised trials [17]. Since most of these drugs have long-lived pharmacological or immunological effects, a protective effect could hypothetically exist after SARS-CoV-2 infection. In our previous analysis of the prevalence of hospital PCR+ cases in rheumatic patients and the general population, a higher prevalence was observed in ts/bDMARD but not csDMARD treated patients [8]. Therefore, our observations regarding ts/bDMARD should be considered with caution because we cannot exclude the possibility of confounding by indication of the therapies in the different included diseases i.e. the preferential use of ts/bDMARD in IA. Larger cohorts of patients treated with these drugs or meta-analysis are warranted to clarify the real impact of ts/bDMARD on COVID-19 susceptibility or severity in rheumatic patients.

Our study has additional limitations. In the time frame where the study was performed in Spain, due to testing shortages, PCR testing selected a population with a certain level of severity since testing of mild cases in the community was not performed. Therefore, we could not introduce the analysis of the full spectrum of the disease, excluding a large proportion of rheumatic patients with a less severe COVID-19.

Since each matched control was also selected in the same date of each case, this was not expected to introduce differences in patients’ profiles. As rheumatic patients and matched controls were selected on the same date, we do not expect to have selection bias. However, fewer patients with radiographic evidence of pneumonia, and a numerical trend towards less respiratory insufficiency and oxygen need were observed in rheumatic patients. This might indicate either a milder expression of lung inflammation in these patients, despite the worse outcome of some groups, or a bias towards more frequent hospital admission and PCR testing due to a higher perception of risk of rheumatic patients.

Since ageing and having an AI/IIMID disease are the most relevant risk factors for severe COVID-19 in patients with rheumatic disease, shared immune-pathogenetic factors that might modify defensive and inflammatory responses need to be identified. Exhaustion of adaptive T-cell responses and increased effector inflammatory responses associated to accumulation of senescent cells, termed inflammaging, have been identified in both situations [18-20]. In experimental models of murine coronavirus infection, biological aging induced by telomeric disfunction, is associated with lethality and higher cytokine responses [21]. In our cohorts, additional differences between rheumatic, including AI/IMID group, and control patients in the expression of COVID-19 disease were not detected. Despite the pro-inflammatory background of AI/IMID patients, the biological response to the infection was similar to that of controls. This suggests that the severity is not necessarily related to quantitative differences in the cytokine (i.e. IL6/CRP) response and additional factors should be searched for.

In conclusion, we found that, among patients with inflammatory rheumatic diseases, having an AI/IMID, aging, and male sex pose a significantly greater risk for poor outcomes, whereas active immunosuppressor therapies do not. This observation should help to tailor prevention measures in specific diagnostic and therapeutic groups of rheumatic patients during this or future coronavirus pandemics.

## Data Availability

Individual de-identified patient data will made available to researchers who provide a reasonable and methodologicaly sound proposal. Proposals should be directed to the corresponding author.

## ACKNOWLEDGEMENTS

We are grateful to Celia Iglesias for valuable help in the preparation of the manuscript.

## COMPETING INTERESTS

None of the authors have competing interests to declare.

## ETHICAL APPROVAL INFORMATION

The study was approved by Comité de Ética de la Investigación del Hospital Universitario 12 de Octubre, CEIm number: 20/160.

## FUNDING INFORMATION

The RIER network was supported by the Fondo de Investigación Sanitaria, Instituto de Salud Carlos III (RD16/0012 RETICS program) and cofinanced by the European Regional Development Fund (FEDER).

## CONTRIBUTORSHIP

JLP, MG and LC take responsibility for the integrity of the data, data analysis and statistical analyses. JLP and LC drafted the manuscript and all authors. All authors participated in acquisition of data, designing the analyses, interpreting the results and critical revision of the manuscript. RIER investigators participated in the design and partially collaborated in acquisition of data. All authors approved the final manuscript.

---

**Table S1.** Autoimmune and Immunomediated diseases (AI/IMID) Diagnostics

| Diagnosis | n |
| --- | --- |
| **Chronic Arthritis** | 136 |
| RA | 65 |
| PsA | 35 |
| SpA | 36 |
| **AI/IMID** | 92 |
| PMR | 24 |
| SLE | 16 |
| SS | 13 |
| SSc | 7 |
| PAPS | 6 |
| Sarcoidosis | 5 |
| GCA | 4 |
| Inflammatory Myopathies | 3 |
| Other Vasculitis | 3 |
| IPAF | 2 |
| Other | 9* |

\* Behcet's, disease, Hidradenitis suppurativa, IgG-4 related disease, Undifferentiated connective tissue disease

## Notes

### Competing Interest Statement

The authors have declared no competing interest.

### Funding Statement

The RIER network was supported by the Fondo de Investigacion Sanitaria, Instituto de Salud Carlos III (RD16/0012 RETICS program) and cofinanced by the European Regional Development Fund (FEDER).

### Author Declarations

The study was approved by Comite de Etica de la Investigacion del Hospital Universitario 12 de Octubre, CEIm number: 20/160.

